# Estimating the Rupture Risk of Large Abdominal Aortic Aneurysms

**DOI:** 10.1101/2025.04.23.25325969

**Authors:** Hataka R Minami, Jayer Chung, Neal R Barshes

**Author notes:** Corresponding author: Neal Barshes, 1 Baylor Plz, Houston, TX 77030.

## Abstract

**Objectives:** Determining the rupture risk of large abdominal aortic aneurysms (AAA) through clinical studies remains elusive. We sought to employ computational methods on existing literature data and briefly report on stratified rupture risk of large AAAs.

**Methods:** We reviewed published English-language literature reporting on rupture risk of large AAAs. Analysis of rupture risk was performed on Microsoft Excel and Visual Basic scripting. Outcome of interest was annual rupture probabilities stratified by aneurysm size (range 5.0/5.5 – 10.0 cm in women/men) and patient sex. Uncertainty in rupture probabilities was considered through probability distributions.

**Results:** For aneurysms sized 5.5 – 7.0 cm, estimated annual rupture probabilities in women were much higher than literature-reported estimates in men, ranging from 6.4% (95% confidence interval: 3.5% – 10.1%) to 54.5% (95% confidence interval: 39.3% – 69.3%). For aneurysms sized 7.5 cm, estimated annual rupture probabilities were 32.4% (95% confidence interval: 26.1% – 39.0%) in men and 64.8% (95% confidence interval: 51.3% – 77.1%) in women. For aneurysms sized 10.0 cm, estimated rupture probabilities increased to near-certainty at 96.6% (95% confidence interval: 94.0% – 98.5%) in men and 98.8% (95% confidence interval: 96.8% – 99.9%) in women.

**Conclusion:** Computational analysis of existing data is feasible to estimate stratified rupture probabilities of large AAAs, based on aneurysm size and patient sex. Further clinical studies are needed to obtain more precise estimates of rupture probabilities.

## Introduction

Rupture risk of abdominal aortic aneurysms (AAAs) has been investigated for decades^1-4^, especially with respect to aneurysm size and patient sex. High-quality data exists for smaller AAAs < 5.0 cm^5, 6^, but large AAAs ≥ 5.0 – 5.5 cm are frequently repaired. This restricts available data on rupture rates^3, 4^. Other limitations in available data include unobserved ruptures and declining AAA rupture rates attributable to reduction in smoking rates, increased penetrance of optimal medical therapy, and increasing utilization of endovascular repair^1-4, 7^. Hence, clinical studies alone may have difficulty estimating rupture risk of large AAAs.

Despite being elusive, estimating rupture risk of large AAAs can be useful. Rupture risk can be weighed against the surgical risk of aneurysm repair, and guide decision-making for individual patients who may have higher surgical risk due to advanced age and/or extensive comorbidities^7^. Formal decision and economic analyses can inform such decision-making, and reliable estimates of AAA rupture risk are fundamental to such analyses^8^. We sought to employ computational methods on existing literature and briefly report on stratified rupture risk of large AAAs, based on aneurysm size and patient sex.

## Methods

To identify studies suitable for computational analysis, we reviewed existing literature reporting on rupture risk of large AAAs (see Supplemental Methods and Supplemental Figure S1). We identified 15 cohort studies^1, 2, 4, 9-20^, 3 randomized controlled trials (RCTs) ^21-23^, and 5 meta-analyses^3, 5, 6, 24, 25^. The extracted study data were summarized in Supplemental Table 1 for cohort studies and randomized controlled trials, and Supplementary Table 2 for meta-analyses. Three out of five meta-analyses focused on large AAAs. These 3 meta-analyses already included 13 out of 15 identified cohort studies, and 1 out of 3 identified RCTs. Among the 2 cohort studies^9, 10^ and 2 RCTs^22, 23^ that were not included in prior meta-analyses, we extracted only 45 additional ruptures out of 394 patients with large AAAs, none of which were stratified by aneurysm size (beyond 5.5 cm) or patient sex.

Performing simple compilation of available rupture data, or even data synthesis via meta-analysis, was unlikely to provide additional meaningful information. Even if performed, this would not provide stratified rupture risk of very large AAAs beyond 7.0 cm, as no studies provided such data (often reported as the size category > 7.0 cm). We therefore selected representative studies for computational analysis. Among the fully eligible studies identified from literature, we selected those with the highest levels of evidence (ordered from highest to lowest: meta-analysis and systematic review, randomized controlled trials, and cohort studies), large sample size, and/or robust estimates of rupture risk. Detailed rationale of study selection were included in Supplemental Tables 1 and 2.

We then performed brief computational analysis of selected studies on Microsoft Excel with Visual Basic scripting. Outcome of interest was annual rupture probabilities stratified by aneurysm size (range 5.0/5.5 – 10.0 cm in women/men) and patient sex. Uncertainty was considered through probability distributions.

For aneurysms sized 5.0 cm in women, we directly used data from a meta-analysis as robust data was available^5^. Rupture data was originally reported in rates (per 1000 person-years), and we converted these rates to probabilities in standard fashion^26^. We included uncertainty by also converting reported 95% confidence intervals from rates to probabilities.

For aneurysms sized 5.5 – 7.0 cm, we directly used the annual cumulative incidence (probabilities) of rupture from the most recent and largest multi-center registry study^4^. In this size range, we did not find suitable systematic reviews or meta-analyses reporting rupture risk stratified by size or sex, and it was already recently attempted without success^24^. Ranges of aneurysm sizes were converted to single sizes for analyses (e.g., 5.5 – 6.0 cm converted to 5.5 cm), and the largest size range > 7.0 cm was converted to 7.0 cm. Rupture probabilities were not separately reported by sex, so we estimated reported probabilities as those of men, because the study population was mostly men (71.2%)^4^.

To estimate corresponding rupture probabilities in women (size range 5.5 – 7.0 cm), we applied a hazard ratio from a separate meta-analysis (HR = 3.76; 95% CI 2.58 – 5.47) ^6^ to take into account that women have approximately 4 times higher risk of rupture above 5.0 cm^2^. The hazard ratio was randomly sampled 1000 times and applied to independently and randomly sampled male rupture probabilities within the same size range (5.5 – 7.0 cm). We used a log-normal distribution for the hazard ratio, and a beta distribution for male rupture probabilities (the latter distribution being standard for underlying probabilities bound between 0 and 1). Independent sampling accounted for combined uncertainty from not only the hazard ratio but also rupture probabilities.

Lastly, there was no data regarding stratified rupture probabilities beyond 7.0 cm. We combined data from RESCAN collaborators for small AAAs (3.0 – 5.0 cm) ^5^, as well as the prior registry study for large AAAs (5.5 – 7.0 cm) ^4^, to perform logistic regression and estimate rupture probabilities beyond 7.0 cm. Regression was performed separately for men and women. The predictor variable was aneurysm size, and the outcome variable was annual rupture probability. We included uncertainty by assuming a normal distribution of the regression coefficients, and randomly sampling from the coefficients 1000 times (based on their standard errors and covariance) to estimate uncertainty ranges of estimated rupture probabilities.

## Results

Annual rupture probabilities were directly estimated from literature up to 7.0 cm in men and 5.0 cm in women (Figure 1 and Table 1). For aneurysms sized 5.5 to 7.0 cm, estimated annual rupture probabilities in women were much higher than those in men, which were previously reported in literature from male-predominant cohorts. At these aneurysm sizes, annual rupture probabilities in women ranged from 6.4% (95% confidence interval: 3.5% - 10.1%) to 54.5% (95% confidence interval: 39.3% - 69.3%).

**Figure 1.**
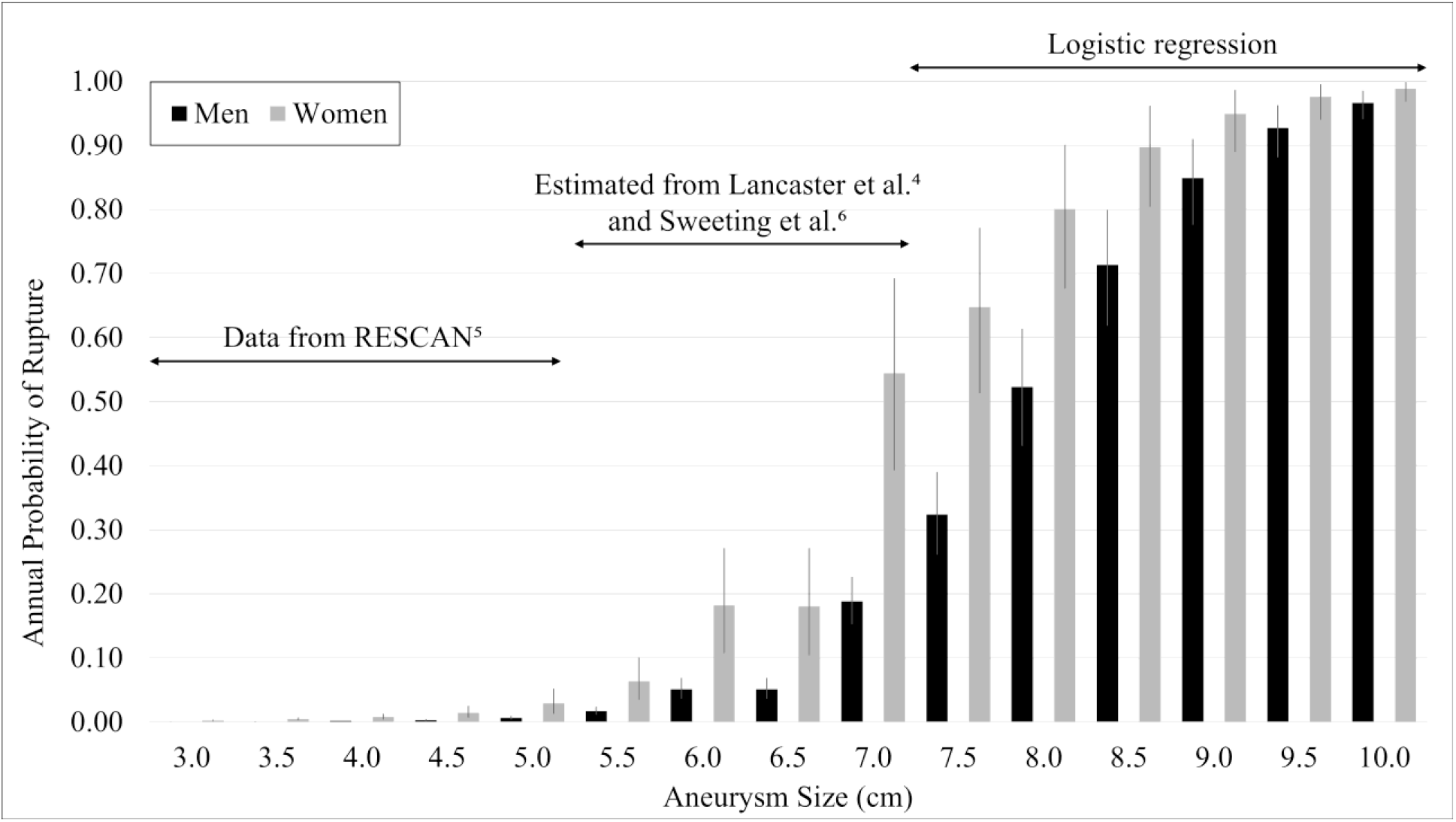
Annual probability of rupture stratified by aneurysm size and sex. Error bars represent 95% confidence intervals.

**Table 1.** Annual Probability of Rupture Stratified by Aneurysm Size and Patient Sex.

| Size | Men | Women |
| --- | --- | --- |
|  | Mean (95% Confidence Interval) | Mean (95% Confidence Interval) |
| **3.0** | 0.0% (0.0% - 0.1%)<sup>5</sup> | 0.2% (0.1% - 0.4%)<sup>5</sup> |
| **3.5** | 0.1% (0.1% - 0.1%)<sup>5</sup> | 0.4% (0.3% - 0.7%)<sup>5</sup> |
| **4.0** | 0.2% (0.1% - 0.2%)<sup>5</sup> | 0.8% (0.4% - 1.4%)<sup>5</sup> |
| **4.5** | 0.3% (0.2% - 0.5%)<sup>5</sup> | 1.5% (0.8% - 2.7%)<sup>5</sup> |
| **5.0** | 0.6% (0.4% - 0.9%)<sup>5</sup> | 2.9% (1.6% - 5.4%)<sup>5</sup> |
| **5.5** | 1.7% (1.2% - 2.5%)<sup>4</sup> | 6.4% (3.5% - 10.1%)* |
| **6.0** | 5.1% (3.6% - 6.9%)<sup>4</sup> | 18.2% (10.7% - 27.1%)* |
| **6.5** | 5.1% (3.6% - 6.9%)<sup>4</sup> | 18.0% (10.4% - 27.1%)* |
| **7.0** | 18.8% (15.3% - 22.6%)<sup>4</sup> | 54.5% (39.3% - 69.3%)* |
| **7.5** | 32.4% (26.1% - 39.0%)** | 64.8% (51.3% - 77.1%)** |
| **8.0** | 52.3% (43.1% - 61.4%)** | 80.1% (67.6% - 90.1%)** |
| **8.5** | 71.3% (61.9% - 79.9%)** | 89.7% (80.5% - 96.2%)** |
| **9.0** | 84.9% (77.6% - 91.0%)** | 94.9% (88.9% - 98.7%)** |
| **9.5** | 92.7% (88.1% - 96.3%)** | 97.6% (94.0% - 99.6%)** |
| **10.0** | 96.6% (94.0% - 98.5%)** | 98.8% (96.8% - 99.9%)** |

\*Estimated with a hazard ratio from Sweeting et al.<sup>6</sup> applied to findings from Lancaster et al.<sup>4</sup> Uncertainty was considered by repeated sampling 1000 times.
\*\*Estimated with logistic regression analysis on existing data for aneurysms sized 3.0 – 7.0 cm. Uncertainty was considered by repeated sampling 1000 times.

When logistic regression analysis was performed using combined data between 3.0 – 7.0 cm, the odds ratio of rupture was 5.26 (95% CI: 4.42 – 6.27) per 1-cm size increase in men, and 5.03 (95% CI: 3.77 – 6.70) per 1-cm size increase in women. For aneurysms sized 7.5 to 10.0 cm, this provided estimated annual rupture probabilities in men from 32.4% (95% confidence interval: 26.1% – 39.0%) to 96.6% (95% confidence interval: 94.0% – 98.5%). Corresponding estimated annual rupture probabilities in women ranged from 64.8% (95% confidence interval: 51.3% - 77.1%) to 98.8% (95% confidence interval: 96.8% - 99.9%).

## Discussion

Estimating the rupture risk of large AAAs remain challenging. In this study, we directly estimated rupture risk from literature up to 7.0 cm in men^4, 5^ and 5.0 cm in women^5, 6^. We did not find suitable literature beyond 7.0 cm in men and 5.0 cm in women. Therefore, we performed computational analysis including application of a hazard ratio for aneurysms sized 5.5 – 7.0 cm in women, and logistic regression for aneurysms sized 7.5 – 10.0 cm in men and women. We believe our analysis integrates the best available evidence regarding rupture risk stratified by aneurysm size and sex. Knowing rupture risk can be particularly useful when weighed against the surgical risk of aneurysm repair for individual patients. For example, patients with higher surgical risk may not derive clinical benefit from repair at standard size thresholds^7^, but may be considered for repair at larger sizes due to higher risk of rupture. Formal decision-making analyses often rely on rupture probabilities stratified by size, and have been previously performed even when these probabilities are very rough estimates. ^8^

Much of existing literature reporting on AAA rupture risks may be outdated and/or be unsuitable for computational analysis. More than two decades ago, Lederle et al. estimated the annual incidence of probable AAA rupture among 198 Veterans (> 90% male smokers) at 9.4%, 10.2%, and 32.5%, respectively, for aneurysms sized 5.5 – 5.9 cm, 6.0 – 6.9 cm, and ≥ 7.0 cm. These rupture probabilities are likely much higher than current estimates (likely due to declining smoking rates, improved medical therapy, and increasing use of endovascular repair^1-4, 7^), including those from Lancaster et al. on which we perform computational analysis^4^. Parkinson et al. conducted a meta-analysis including 1541 patients unfit for surgery (mostly men), and found the annual incidence of AAA rupture at 3.5%, 4.1%, and 6.3%, respectively, for AAAs sized 5.5 – 6.0 cm, 6.1 – 7.0 cm, and > 7.0 cm. However, this study included 9 out of 11 studies from two decades ago (starting from 1998), and did not stratify rupture probabilities by sex. Leone et al. recently performed another meta-analysis (2023) and reported on a pooled rupture probability of 25.7% for AAAs > 5.5 cm, over a follow-up of approximately 2 years^24^. Although including more recent studies (3 out of 4 within the last decade), they could not fully stratify by aneurysm size or patient sex due to scarcely reported data.

Interpretation of rupture data must be made with caution, especially for large aneurysms where patients may be at competing risks of rupture and repair. For example, Lancaster et al. accounted for competing risks by using a competing risk model^4^, which prevents over-estimation of rupture incidence compared to the standard Kaplan-Meier function^27^. However, the competing risk model can still underestimate rupture probabilities of aneurysms that remain unrepaired. Originally reported cumulative incidences of rupture for aneurysms sized 5.5 – 6.0 cm were 1.7%/2.5%/3.1% at 12/24/36-month follow-ups. When these estimates were converted to annual probabilities (using rate conversion as an intermediary step^26^), we obtained annual rupture probabilities that decrease over time at 1.7%/1.3%/1.0% per year, respectively. These estimates represent rupture probabilities where aneurysm repair is occurring, and rupture probabilities are correctly adjusted downward. Importantly, we aim to estimate rupture probabilities where aneurysm repair is not occurring. True rupture probabilities of unrepaired AAAs likely stay the same every year given unchanged patient demographics, or even increase with aneurysm growth. In our study, we used the 12-month cumulative incidence (rather than 36 months) from Lancaster et al. to minimize any underestimate of rupture probabilities.

Our study has limitations inherent to computational analysis. First, we estimated rupture probabilities in women separately from men, based on the understanding that women have higher risk of rupture.^6^ The RESCAN collaborators identified a 4-fold higher risk of rupture for small AAAs sized 3.0 – 5.0 cm, and this was surprisingly consistent across the entire size range^5^. Sweeting et al. confirmed an approximately 4-fold higher risk of rupture for small AAAs^6^. Brown et al. showed that women have similar 4-fold higher rupture risk beyond 5.0 cm, at least up to 6.0 cm^2^. It is unclear whether the same 4-fold higher risk applies up to 7.0 cm, but we included uncertainty through a probability distribution and repeated sampling. Lastly, we used logistic regression to extrapolate rupture probabilities beyond 7.0 cm. Regression analysis is uncertain beyond the original data (3.0 – 7.0 cm) that was used to perform the analysis. However, rupture probabilities are expected to continue increasing with size. Our model predicts narrowing confidence intervals, or increasing certainty of rupture, as probabilities reach near-certainty for very large aneurysms around 10.0 cm.

Despite these limitations, obtaining stratified rupture probabilities of large AAAs based on size and sex can be worthwhile. More precise estimates of rupture probabilities, especially for AAAs > 7.0 cm, can be provided with prospective registries or randomized controlled trials involving observation of unrepaired aneurysms. We hope that our brief analysis highlights the poorly characterized aspects of the natural history of large AAAs, and that our estimates are informative to clinicians and spur future research.

## Conclusion

Computational analysis of existing data is feasible to estimate stratified rupture probabilities of large AAAs, based on aneurysm size and patient sex.

Further clinical studies are needed to obtain more precise estimates of rupture probabilities.

## Data Availability

All data produced in the present study are available upon reasonable request to the authors.

## Notes

### Competing Interest Statement

The authors have declared no competing interest.

### Funding Statement

This study did not receive any funding.

