## Supplemental Material for "Estimating the Rupture Risk of Large Abdominal Aortic Aneurysms"

**Supplemental Methods**

We comprehensively searched MEDLINE via PubMed, Embase, and Cochrane Central Register of Controlled Trials using the exploded medical subject headings “abdominal aortic aneurysm” and “aortic rupture.” We filtered for studies with English-language restrictions and those published after January 1, 2000. We also reviewed bibliographies and the similar articles function on PubMed to identify additional studies that may be eligible. The search was closed on December 28, 2024.

Studies were eligible for inclusion with the following criteria: (1) men or women ≥ 18 years old; (2) men with unrepaired large abdominal aortic aneurysms (AAAs) and index (initial) sizes ≥ 5.5 cm and/or women with unrepaired large AAAs and index sizes ≥ 5.0 cm; (3) undergoing screening or surveillance; (4) availability of ultrasound, computed tomography, or magnetic resonance imaging; and (5) reporting rupture risk. Studies were excluded with the following criteria: (1) AAA sizes limited to < 5.5 in men and < 5.0 cm in women; (2) not reporting rupture risk; (3) reporting mostly on aneurysm repair; (4) reporting mostly thoracic or thoracoabdominal aortic aneurysms; (5) sample sizes ≤ 10 participants; and (6) publications in the form of abstracts, posters, case reports, editorials, letters, replies, commentary, and discussion/debate. Systematic reviews and meta-analyses were considered eligible if they included studies meeting the above inclusion and exclusion criteria. The study title followed by the abstract was reviewed for inclusion and exclusion criteria. Studies with screened abstracts underwent full-text review.

Among fully eligible studies, we extracted study data (see Supplemental Tables 1 and 2) including the following: study authors, study country, study design, study year, AAA size, number of large AAAs, rupture data, and rationale for study selection for computational analysis.

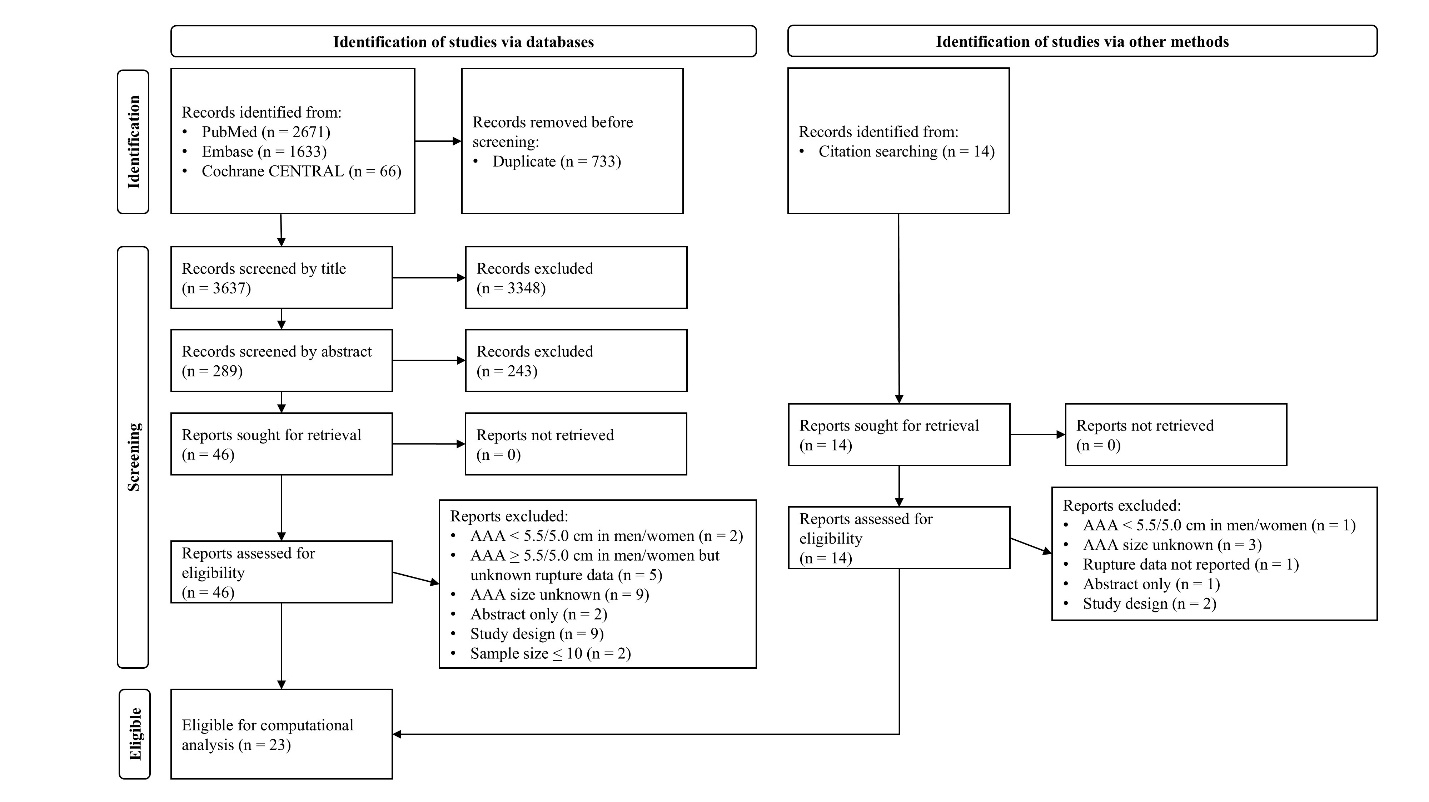
Supplemental Figure S1. Flowchart of eligible studies.

**Supplemental Table S1. Data Extraction Sheet of Cohort Studies and Randomized Controlled Trials**

| **Study Authors** | **Study Country** | **Study Design** | **Study Year** | **AAA Size** | **Sample Size** | **Description of Rupture Data** | **Study Selection for Computational Analysis and Rationale** |
| --- | --- | --- | --- | --- | --- | --- | --- |
| Talvitie et al. ^9^ | Sweden, Norway, and Austria | Retrospective multi-center cohort study | 2023 | Women: median 5.0 cm | Women: 51 | Women: 9 ruptures over median 2.0 years of follow-up | Not selected due to small sample size. |
| Lancaster et al. ^4^ | United States | Retrospective multi-center cohort study | 2022 | Men: ≥ 5.5 cm  Women: ≥ 5.0 cm | Men and women: 3248 (71.2% men) | Annual cumulative incidence of confirmed and probable rupture:  Women:  5.0 – 5.4 cm: 2.7% (1.6 - 4.2%)  Men and women:  5.5 – 6.0 cm: 1.7% (1.2 - 2.5%)  6.1 – 7.0 cm: 5.1% (3.6 - 6.9%)  > 7.0 cm: 18.8% (15.3 - 22.6%) | Selected due to largest sample size and being conducted within the last 10 years. |
| Gallitto et al. ^10^ | Italy | Retrospective single-center cohort study | 2020 | Men and women: average 56 mm (juxtarenal/pararenal) | Men and women: 75 | Single rupture (unknown sex) after 45 days of customized graft manufacturing | Not selected due to small sample size. |
| Elmallah et al. ^11^ | Ireland | Retrospective single-center cohort study | 2019 | Men and women: > 5.5 cm | Men: 54  Women: 22 | Men and women: 16 ruptures over average 24.9 months of follow-up | Not selected due to small sample size. |
| EVAR-2^21^ | United Kingdom | Randomized controlled trial (EVAR-2) | 2017 | Men and women: ≥ 5.5 cm | Men and women: 207 | Men and women: 53 ruptures over average 4.2 years of follow-up | Not selected due to lack of size granularity and relatively small sample size. |
| Scott et al. ^12^ | United Kingdom | Retrospective single-center cohort study | 2016 | Men and women: > 5.5 cm | Men:  5.5 – 5.9 cm: 42  6.0 – 6.9 cm: 47  ≥ 7.0 cm: 26  Women:  5.5 – 5.9 cm: 7  6.0 – 6.9 cm: 11  ≥ 7.0 cm: 5 | Men and women: 37 ruptures over median 27 months of follow-up | Not selected due to relatively small sample size. |
| Lim et al. ^13^ | United Kingdom | Retrospective single-center cohort study | 2015 | Men: > 5.4 cm | Men: 59 | Men: 10 ruptures over median 24.1 months of follow-up | Not selected due to small sample size. |
| Al-Thani et al. ^14^ | Qatar | Retrospective single-center cohort study | 2014 | Men and women: ≥ 5.5 cm | Men and women:  5.5 – 6.9 cm: 22  ≥ 7.0 cm: 14 | Men and women:  5.0 – 5.9 cm: 2 ruptures  6.0 – 6.9: 2 ruptures  ≥ 7.0 cm: 6 ruptures  Over 3 years of follow-up | Not selected due to small sample size. |
| Western et al. ^16^ | United Kingdom | Retrospective single-center cohort study | 2013 | Men: > 5.5 cm  Women: > 5.0 cm | Men: 55  Women: 17 | Men and women:  5.1 – 6.0 cm: 5 ruptures out of 45 patients  6.1 – 7.0 cm: 4 ruptures out of 20 patients  > 7.0 cm: 3 ruptures out of 7 patients  Over a study period of approximately 6 years | Not selected due to small sample size. |
| Noronen et al. ^15^ | Finland | Retrospective single-center cohort study | 2013 | Men and women: ≥ 5.5 cm | Men and women:  5.5 – 6.0 cm: 74  6.1 – 7.0 cm: 57  > 7.0 cm: 23 | Men and women:  5.5 – 6.0 cm: 24 ruptures  6.1 – 7.0 cm: 22 ruptures  > 7.0 cm: 10 ruptures  Over a median follow-up of 19.0 months | Not selected due to relatively small sample size. |
| Scott et al. ^22^ | United Kingdom | Sub-analysis of a randomized controlled trial (MAAS) | 2005 | Men: ≥ 5.5 cm | Men: 168 | Men: 15 ruptures  Rate: 140 (78 – 231) per 1000 person-years | Not selected due to relatively small sample size. |
| Aziz et al. ^17^ | New Zealand | Retrospective single-center cohort study | 2004 | Men and women: ≥ 5.0 cm | Men and women:  5.0 – 5.9 cm: 58  ≥ 6.0 cm: 53 | Men and women:  5.0 – 5.9 cm: 5 ruptures over median follow-up of 16.8 months  ≥ 6.0 cm: 22 ruptures over median follow-up of 14.1 months  Overall ≥ 5.0 cm: annual cumulative incidence of rupture = 13% | Not selected due to relatively small sample size. |
| Tambyraja^18^ | United Kingdom | Retrospective single-center cohort study | 2003 | Men and women: ≥ 5.5 cm | Men and women:  5.5 – 5.9 cm: 16  6.0 – 6.9 cm: 28  ≥ 7.0 cm: 18 | Men and women:  5.5 – 5.9 cm: 3 AAA deaths over median follow-up of 19 months  6.0 – 6.9 cm: 9 AAA deaths over median follow-up of 21 months  ≥ 7.0 cm: 6 AAA deaths over median follow-up of 8 months | Not selected due to small sample size. |
| Brown et al. ^2^ | Canada | Prospective single-center cohort study | 2003 | Men and women: ≥ 5.0 cm | Men and women: 400 | Men and women: 50 ruptures over 982 patient-years of follow-up | Not selected due to lack of size granularity and relatively small sample size. |
| Tanquilut^19^ | United States | Retrospective single-center cohort study | 2002 | Men and women: ≥ 5.6 cm | Men and women: 19 | Men and women: 3 ruptures over average 29 months of follow-up | Not selected due to small sample size. |
| Lederle et al. ^1^ | United States | Prospective multi-center cohort study | 2002 | Men and women: ≥ 5.5 cm | Men and women:  5.5 – 5.9 cm: 61  6.0 – 6.9 cm: 85  ≥ 7.0 cm: 52  (Note: only 1 woman) | Annual cumulative incidence of definite and probable rupture:  Men and women:  5.5 – 5.9 cm: 9.4%  6.0 – 6.9 cm: 10.2%  > 7.0 cm: 32.5% | Not selected due to relatively small sample size and likely lower contemporary rupture rates compared to those reported here. |
| Conway et al. ^20^ | United Kingdom | Retrospective single-center cohort study | 2001 | Men and women: ≥ 5.5 cm | Men:  5.5 – 5.9 cm: 14  6.0 – 7.0 cm: 39  > 7.0 cm: 17  Women:  5.5 – 5.9 cm: 9  6.0 – 7.0 cm: 23  > 7.0 cm: 4 | Men and women:  5.5 – 5.9 cm: 5 ruptures  6.0 – 7.0 cm: 21 ruptures  > 7.0 cm: 11 ruptures | Not selected due to relatively small sample size. |
| Powell et al. ^23^ | United Kingdom | Sub-analysis of a randomized controlled trial (UK Small Aneurysm Trial) | 2001 | Men and women: 5.6 – 9.7 cm | Men and women: 100 | Men and women: 20 ruptures  Rupture rate: 27.8 per 100 person-years  Hazard ratio for rupture: 2.94 (2.49 – 3.48) per 1.0 cm increase in size | Not selected due to lack of size granularity and relatively small sample size. |

**Supplemental Table S2. Data Extraction Sheet of Meta-Analyses**

| **Study Authors** | **Study Country** | **Study Design** | **Study Year** | **Aneurysm Size** | **Sample Size** | **Description of Rupture Data** | **Study Selection for Computational Analysis and Rationale** |
| --- | --- | --- | --- | --- | --- | --- | --- |
| Leone et al. ^24^ | Denmark | Meta-analysis | 2023 | Men and women: > 5.5 cm | Men and women: 427 | Men and women: 119 ruptures with unknown pooled follow-up duration | Not selected due to lack of size granularity and relatively small sample size. |
| Parkinson et al. ^3^ | United Kingdom | Meta-analysis | 2015 | Men and women: ≥ 5.5 cm | Men and women: 1514 (mostly male but unclear proportion) | Annual cumulative rupture rates:  Men and women:  5.5 – 6.0 cm: 3.5% (-1.6 – 8.7%)  6.1 – 7.0 cm: 4.1% (-0.7% – 9.0%)  > 7.0 cm: 6.3% (-1.8 – 14.3%) | Not selected due to:  1. Most (9 out of 11) studies were published > 15 years ago.  2. Despite being a meta-analysis, sample size was still smaller than Lancaster et al. |
| RESCAN Collaborators^5^ | United Kingdom | Meta-analysis | 2013 | Women: 5.0 – 5.4 cm | Women: 1743 for size range 3.0 – 5.4 cm (not reported for 5.0 – 5.4 cm) | Women:  5.0 cm: rupture rate 29.7 (15.9 – 55.4) per 1000 person-years  Women: 4-fold higher risk of compare compared to men. | Selected due to being the most robust single-size estimate for rupture at 5.0 cm in women.  Notably, this rupture rate of 29.7 per 1000 person-years can be converted to annual rupture probability (2.93%), which is very close to the estimate from Lancaster et al. (2.7%). |
| Sweeting et al. ^6^ | United Kingdom | Meta-analysis | 2012 | Women: 5.0 – 5.4 cm | Women: ~ 1744 for size range 3.0 – 5.4 cm (not reported for 5.0 – 5.4 cm) | Hazard ratio for rupture being a woman: 3.76 (2.58 – 5.47) | Selected due to being the most robust estimate for hazard ratio for rupture being a woman. |
| Powell et al. ^25^ | United Kingdom | Meta-analysis | 2008 | Men and women: ≥ 5.0 cm | Men and women:  5.0 – 5.9 cm: 232  ≥ 6.0 cm: 301 | Men and women: 163 ruptures  5.0 – 5.9 cm: rupture rate 10.3 (7.5 – 14.3) per 100 person-years  ≥ 6.0 cm: rupture rate 27.0 (21.1 – 34.7) per 100 person-years  Overall: rate 18.2 (13.7 – 24.1) per 100 person-years | Not selected due to being published > 10 years ago, and relatively small sample size. |
